# GLP-1 receptor agonists and the risk of acute pancreatitis: a living systematic review and meta-analysis

**DOI:** 10.64898/2026.03.19.26348844

**Authors:** Liam Bakker, Thomas Caganek, Amrit Rooprai, Samuel Hume

## Abstract

**Background:** There are concerns that glucagon-like peptide-1 receptor agonists (GLP1s) increase the risk of acute pancreatitis.

**Methods:** We searched Medline and clinicaltrials.gov in March 2026 for placebo-controlled randomized trials of semaglutide and tirzepatide reporting the incidence of acute pancreatitis, according to a pre-specified protocol (PROSPERO ID 1346039). The primary analysis used a fixed-effect Mantel–Haenszel odds ratio. Heterogeneity was assessed using Cochran’s Q and I-squared.

**Results:** 1635 studies were found, 31 placebo-controlled trials of which were included, totalling 40,274 patients. There were 59 acute pancreatitis events in GLP1 groups (of 22,841 patients) and 50 in placebo groups (of 17,433 patients). The pooled fixed-effect Mantel–Haenszel odds ratio was 0.99 (95% confidence interval 0.67 to 1.45; p=0.95). Active-treatment exposure totaled 51,346 patient-years, including 47,749 patient-years for semaglutide and 3,598 patient-years for tirzepatide. Sensitivity analysis for risk ratio, and subgroup analysis divided by drug class, disease focus, or dose, did not reveal any significant differences.

**Conclusions:** Semaglutide and tirzepatide were not associated with an increased risk of acute pancreatitis versus placebo. These findings are reassuring, but small differences in risk cannot be fully excluded given the rarity of events. Given the rapidly-evolving nature of this field and the importance of the dataset, this review will be updated as further randomized trials are published.

## 1. Introduction

GLP1s have transformed the management of obesity and type 2 diabetes, and are increasingly used across a broader spectrum, including for cardiovascular prevention, chronic kidney disease, heart failure, and metabolic dysfunction-associated steatohepatitis^1^. Semaglutide and tirzepatide are now the first and second-top selling drugs globally, and over 95% of the GLP1-receiving population are taking subcutaneous formulations of these drugs^2^.

One safety question that has persisted is the possibility of acute pancreatitis. The main causes of acute pancreatitis are gallstones, followed by excessive alcohol consumption, but drugs are among the potential causes too^3^. Pancreatitis is inflammation of the pancreas; it presents with abdominal pain, is typically treated supportively (or with surgery, if caused by gallstones). Complications include organ failure, pancreatic necrosis, chronic pancreatitis, and pancreatic pseudocysts; the case fatality rate is 1–7%^4^.

This concern has biological plausibility, given hypothesized direct pancreatic effects of GLP1s^5^ and a plausible indirect pathway through gallstone disease^6^. Regulators around the world continue to recognise acute pancreatitis as a potential safety signal.

An observational study found an increased risk of pancreatitis with GLP1s (pooled dulaglutide, lixisenatide, exenatide, liraglutide, and semaglutide^7^) and a meta-analysis of randomized trials (pooled dulaglutide, exenatide, liraglutide, semaglutide, beinaglutide, retatrutide, or tirzepatide) found a small increased risk with GLP1s^8^. Observational studies are prone to bias, however, and the meta-analysis now misses many of the updated studies, and does not focus (or provide a sensitivity analysis) for the GLP1s most relevant globally today: semaglutide and tirzepatide.

We therefore conducted an up-to-date meta-analysis of placebo-controlled randomized trials focussed on semaglutide and tirzepatide, to evaluate the risk of acute pancreatitis. Given the rapidly-evolving nature of this field, we asked whether there is any causal relationship with GLP1s and acute pancreatitis.

## 2. Methods

### 2.1. Study design

We performed a trial-level meta-analysis of randomized, placebo-controlled trials evaluating semaglutide or tirzepatide and reporting acute pancreatitis events. The systematic review was performed according to the PRISMA guidelines, and registered on PROSPERO (ID: 1346039).

### 2.2. Search strategy

We searched Pubmed and clinicaltrials.gov in March 2026 for

> ((“semaglutide”[tiab] OR “tirzepatide”[tiab]) AND (randomized controlled trial[pt] OR controlled clinical trial[pt] OR randomized[tiab] OR randomised[tiab] OR randomly[tiab] OR trial[tiab]))

### 2.3. Screening

Studies were screened by title, abstract, and full text, according to the inclusion and exclusion criteria.

### 2.4. Inclusion criteria

Population: any; Intervention: subcutaneous tirzepatide or subcutaneous semaglutide, tested in a phase 3 trial; Comparator: placebo; Outcome: acute pancreatitis.

### 2.5. Exclusion criteria

#### Interventions

Active-comparator-only trials; trials with pre-randomization exposure to a GLP-1 receptor agonist judged likely to contaminate treatment comparison; trials that do not report acute pancreatitis as a safety outcome.

#### Study Types

Case Reports, Editorials, Letters To The Editor, Conference Abstracts Only, Narrative Reviews, Protocols, Meta-analysis, Analyses Of Multiple Trials, Systematic Reviews, Phase 2 Trials, post hoc analyses without independent randomized comparison data.

#### Participants

Animal Studies, In Vitro Studies.

### 2.6. Data extraction

For each trial, we extracted the study identifier, drug, number of acute pancreatitis events in the active-treatment arm, total number of participants in the active-treatment arm, number of acute pancreatitis events in the placebo arm, and total number of participants in the placebo arm. Trials with zero events in both arms were retained in the descriptive table but were not estimable for study-level odds ratios and did not contribute to the pooled Mantel–Haenszel relative-effect estimate.

### 2.7. Statistical analysis

The primary analysis used a fixed-effect Mantel–Haenszel model to pool odds ratios with 95% confidence intervals. This approach was selected because acute pancreatitis was a rare event and because the included placebo-controlled trial comparisons showed little between-study heterogeneity. Study-level odds ratios were presented for estimable studies. Double-zero studies were labeled not estimable.

Heterogeneity was assessed using Cochran’s Q statistic and I-squared. Statistical significance for the pooled effect was assessed using the Z test. A two-sided p value of less than 0.05 was considered statistically significant.

### 2.8. Use of AI

We used ScholaraAI, a platform we developed and validated^9^, to perform the first search, screening, data extraction, and analysis. We performed these steps again manually – independently – and a third researcher acted as tiebreaker, when required.

## 3. Results

### 3.1. Included trials and participants

We found 1635 studies in our search, 30 of which^10–39^ were included (a total of 31 randomized trials) (Figure 1). The trials and their characteristics are listed in Supplementary Table 1. These comprised 22,841 participants randomized to semaglutide or tirzepatide and 17,433 participants randomized to placebo.

**Figure 1.**
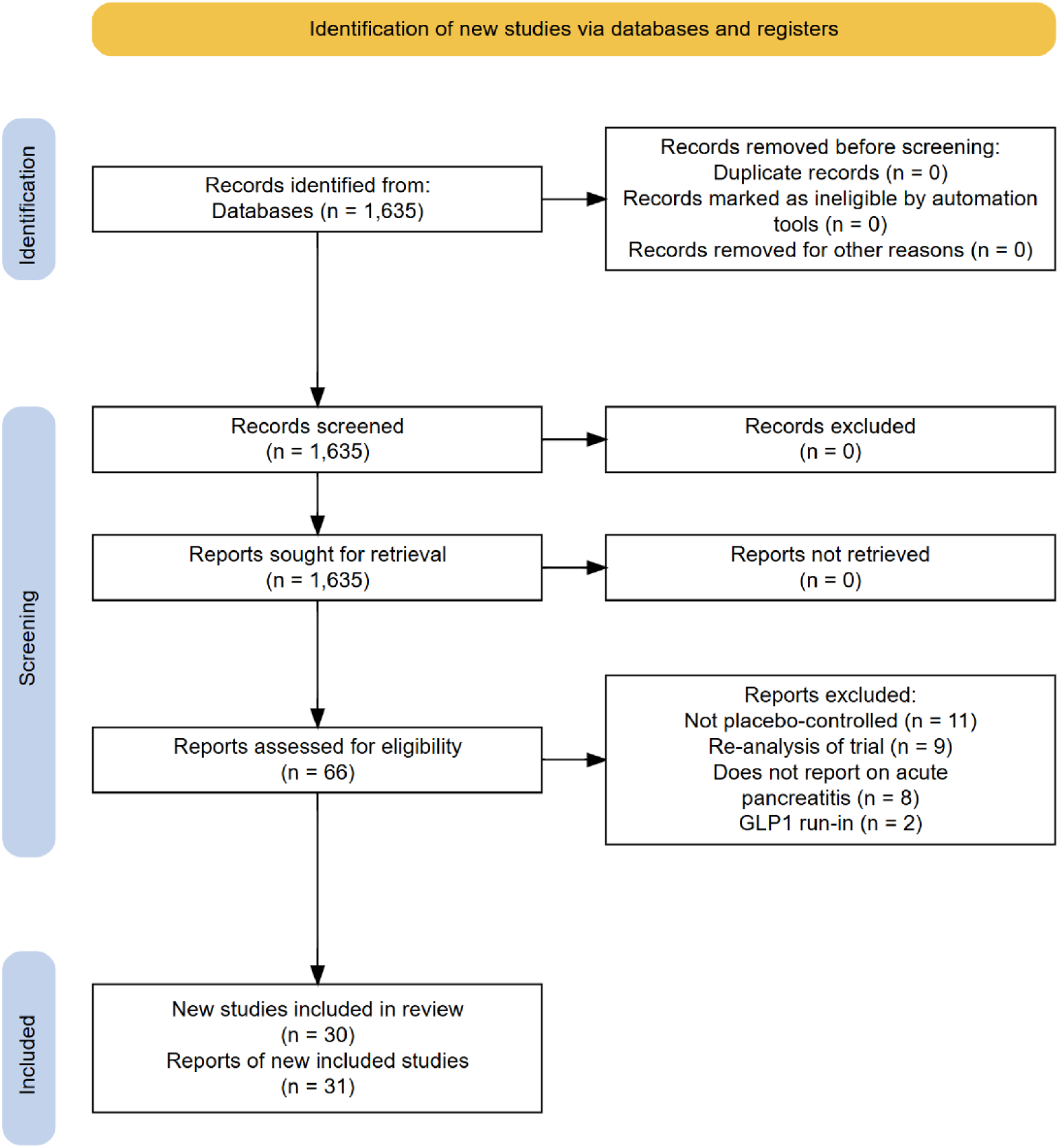
PRISMA flow chart.

### 3.2. Acute pancreatitis events

Across all included trials, there were 59 acute pancreatitis events among 22,841 participants in the active-treatment groups and 50 events among 17,433 participants in the placebo groups (Figure 2). The pooled fixed-effect Mantel–Haenszel odds ratio for acute pancreatitis was 0.99 (95% confidence interval 0.67 to 1.45; p=0.95), indicating no evidence of an increased risk with semaglutide or tirzepatide compared with placebo.

**Figure 2.**
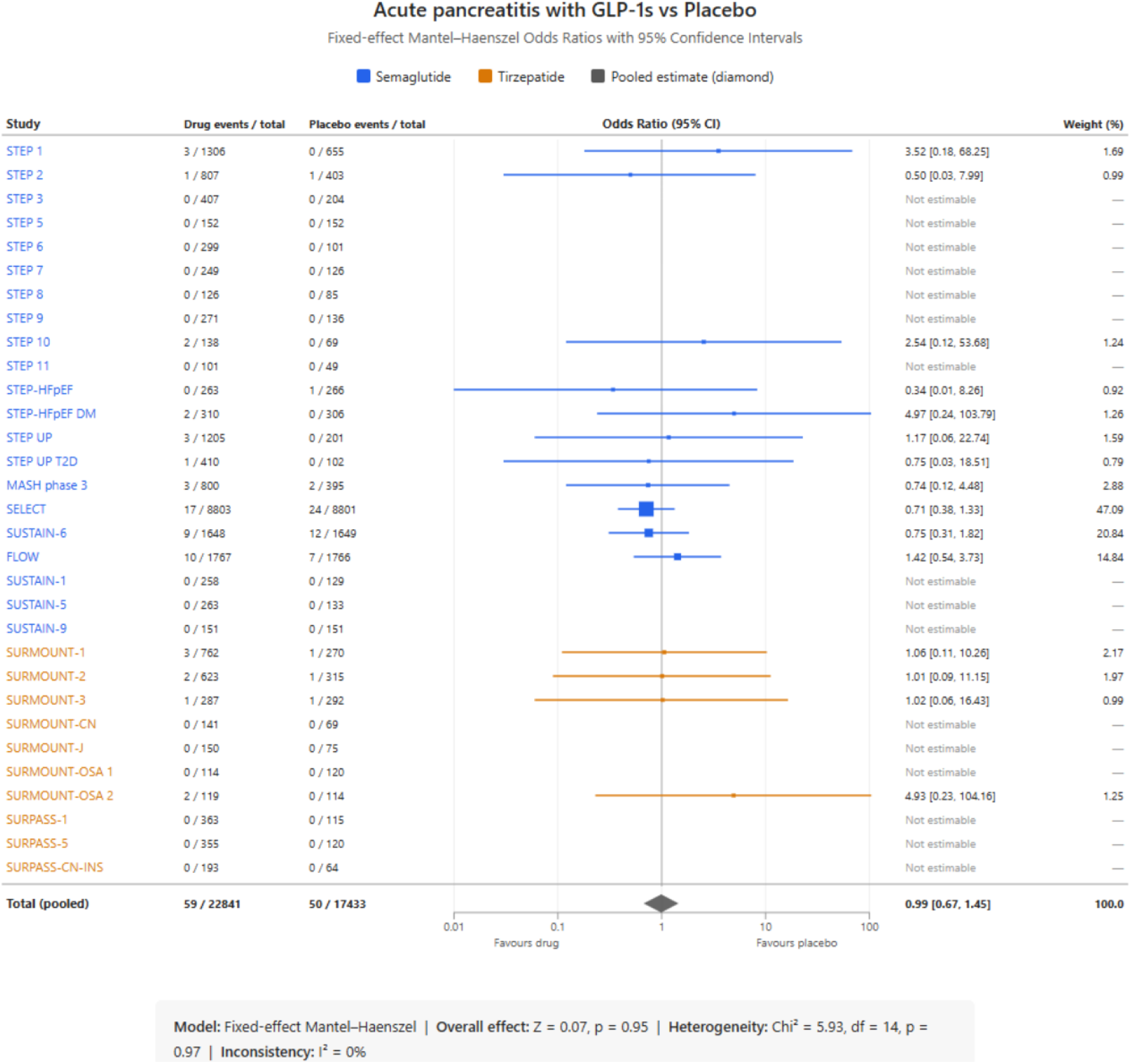
Meta-analysis of 31 GLP1 vs. placebo randomized trials.

Heterogeneity was negligible, with Cochran’s Q of 5.93 on 14 degrees of freedom (p=0.97) and an I-squared of 0%. The pooled estimate was therefore highly consistent across the estimable studies.

The greatest statistical weight came from the large semaglutide outcome trials, particularly SELECT and FLOW, with additional important contribution from SUSTAIN-6. Smaller obesity and diabetes trials contributed less weight, and many had no pancreatitis events in either group.

### 3.3. Sensitivity analysis

Because acute pancreatitis was a rare event, we performed a sensitivity analysis for risk ratio, to check that the conclusion did not depend on the choice of effect measure. The risk ratio was 0.99 (0.67–1.46) (Supplementary Figure 1).

### 3.4. Subgroup analysis

#### Semaglutide or tirzepatide alone

To determine whether there was a significant difference with either semaglutide or tirzepatide alone, we performed subgroup analyses. For semaglutide, with 21 trials, the odds ratio was 0.94 (0.63–1.41) (Supplementary Figure 2). For tirzepatide, with 10 trials, the odds ratio was 1.55 (0.41–5.84) (Supplementary Figure 3), suggesting no significant difference with either semaglutide or tirzepatide alone.

#### By highest-dose only

To determine whether there was a significant difference with those on the highest dose of drug, we performed subgroup analysis for this. There was no significant difference (Supplementary Figure 4).

#### By disease class

To determine whether there was a significant difference with different disease state, we performed subgroup analysis in patients with obesity/overweight (Supplementary Figure 5), type 2 diabetes (Supplementary Figure 6), heart failure (Supplementary Figure 7), chronic kidney disease (Supplementary Figure 8), cardiovascular disease (Supplementary Figure 9), or metabolic dysfunction-associated steatohepatitis (Supplementary Figure 10). We found no significant difference in any subgroup.

### 3.5. Publication bias

Publication bias was assessed visually by inspection of a funnel plot of log-transformed odds ratios against their standard errors, and formally by Egger’s regression test and Begg and Mazumdar’s rank correlation test (Supplementary Figure 11). Of the 31 included studies, 15 had estimable effect sizes and contributed to the assessment; the remaining 16 reported zero events in both arms. The funnel plot appeared broadly symmetrical. Egger’s regression test did not indicate statistically significant asymmetry (intercept = 0.49, SE 0.27; t = 1.80, df = 13, p = 0.095), nor did Begg’s test (Kendall’s *τ* = 0.23, z = 1.14, p = 0.255). These findings suggest no strong evidence of publication bias, although the power of both tests is limited given the small number of informative studies and the rarity of events.

## 4. Discussion

In this meta-analysis of placebo-controlled randomized trials, semaglutide and tirzepatide were not associated with an increased risk of acute pancreatitis compared with placebo. The pooled effect estimate was essentially null, the confidence interval crossed unity. Taken together, these findings provide reassurance that a clear pancreatitis safety signal is not evident in the randomized placebo-controlled data currently available for these agents.

These findings should be interpreted in the context of event rarity. Acute pancreatitis was uncommon in both active-treatment and placebo groups, and many trials had zero events in both arms. This is clinically reassuring, but it also means the analysis is fundamentally a rare-event meta-analysis. Accordingly, while our findings argue against a substantial excess risk, the confidence interval remains compatible with modest harm or modest benefit, and very small differences in risk cannot be excluded.

There are strengths to this dataset. First, it is the most up-to-date analysis of causative data on the link between the modern GLP1s used by most patients today. Second, it is a comprehensive systematic review, containing all of the randomized data that report on this important adverse event. Third, the dataset is living: this is a rapidly-evolving field, and as more trials are published – and with newer GLP1s – we will update and re-publish the data.

This study has limitations. First, the analysis was based on trial-level rather than patient-level data. Second, because the event is rare, we used the fixed-effect Mantel–Haenszel approach, but alternative rare-event methods may produce slightly different numerical estimates. Third, our synthesis was designed around placebo-controlled comparisons and therefore does not address comparative risk versus other active therapies. Fourth, we did not include next-generation GLP1s, that are not yet approved (oral GLP1s, triple agonists like Retatrutide, or GLP1-amylin agonists like Cagrisema).

Overall, the present data support the conclusion that semaglutide and tirzepatide are not associated with a detectable increase in acute pancreatitis risk versus placebo in randomized trials. Given the rapidly-evolving nature of this field, we will update this work when more data are available.

## Data Availability

Analysis of public data

