## Supplementary data for "GLP-1 receptor agonists and the risk of acute pancreatitis: a living systematic review and meta-analysis"

Supplementary Table 1: Characteristics of included studies

Supplementary Figure 1: Sensitivity analysis for risk ratio

Supplementary Figure 2: Subgroup analysis for semaglutide

Supplementary Figure 3: Subgroup analysis for tirzepatide

Supplementary Figure 4: Subgroup analysis by dose

Supplementary Figure 5: Subgroup analysis by overweight/obesity

Supplementary Figure 6: Subgroup analysis by type 2 diabetes

Supplementary Figure 7: Subgroup analysis by heart failure

Supplementary Figure 8: Subgroup analysis by chronic kidney disease

Supplementary Figure 9: Subgroup analysis by cardiovascular disease

Supplementary Figure 10: Subgroup analysis by metabolic dysfunction-associated steatohepatitis

Supplementary Figure 11: Publication bias funnel plot

Supplementary table 1: Characteristics of included studies

| Trial | Drug | Population studied | Active dose(s) in trial | Comparator | Treatment duration | Total randomized | Disease class |
|---|---|---|---|---|---|---|---|
| STEP 1 | Semaglutide | Adults with overweight or obesity, without diabetes | Semaglutide 2.4 mg once weekly | Placebo | 68 weeks | 1961 | Obesity/overweight |
| STEP 2 | Semaglutide | Adults with overweight or obesity and type 2 diabetes mellitus | Semaglutide 2.4 mg and 1.0 mg once weekly | Placebo | 68 weeks | 1210 | Obesity/overweight + type 2 diabetes mellitus |
| STEP 3 | Semaglutide | Adults with overweight or obesity, without diabetes; intensive behavioural therapy background | Semaglutide 2.4 mg once weekly | Placebo | 68 weeks | 611 | Obesity/overweight |
| STEP 5 | Semaglutide | Adults with overweight or obesity, without diabetes | Semaglutide 2.4 mg once weekly | Placebo | 104 weeks | 304 | Obesity/overweight |
| STEP 6 | Semaglutide | East Asian adults with obesity, with or without type 2 diabetes mellitus | Semaglutide 2.4 mg and 1.7 mg once weekly | Placebo | 68 weeks | 400 | Obesity/overweight |
| STEP 7 | Semaglutide | Predominantly East Asian adults with overweight or obesity, with or without type 2 diabetes mellitus | Semaglutide 2.4 mg once weekly | Placebo | 44 weeks | 375 | Obesity/overweight |
| STEP 8 | Semaglutide | Adults with overweight or obesity, without diabetes | Semaglutide 2.4 mg once weekly | Placebo and liraglutide active comparator | 68 weeks | 211 | Obesity/overweight |
| STEP 9 | Semaglutide | Adults with overweight or obesity and type 2 diabetes mellitus | Semaglutide 2.4 mg once weekly | Placebo | 68 weeks | 407 | Obesity/overweight + type 2 diabetes mellitus |
| STEP 10 | Semaglutide | Adults with obesity after prior weight loss | Semaglutide 2.4 mg once weekly | Placebo | 68 weeks | 207 | Obesity/overweight |
| STEP 11 | Semaglutide | Adults with obesity and type 2 diabetes mellitus in East Asia | Semaglutide 2.4 mg once weekly | Placebo | 68 weeks | 150 | Obesity/overweight + type 2 diabetes mellitus |
| STEP-HF pEF | Semaglutide | Patients with obesity-related heart failure with preserved ejection fraction | Semaglutide 2.4 mg once weekly | Placebo | 52 weeks | 529 | Heart failure with preserved ejection fraction |
| STEP-HF pEF DM | Semaglutide | Patients with obesity-related heart failure with preserved ejection fraction and type 2 diabetes mellitus | Semaglutide 2.4 mg once weekly | Placebo | 52 weeks | 616 | Heart failure with preserved ejection fraction |
| STEP UP | Semaglutide | Adults with obesity, without | Semaglutide | Placebo | 72 weeks | 1407 | Obesity/overweight |

| | | | | | | | |
|---|---|---|---|---|---|---|---|
| | | diabetes | 7.2 mg and 2.4 mg once weekly | | | | |
| STEP UP T2D | Semaglutide | Adults with obesity and type 2 diabetes mellitus | Semaglutide 7.2 mg and 2.4 mg once weekly | Placebo | 72 weeks | 512 | Obesity/overweight + type 2 diabetes mellitus |
| MASH phase 3 (ESSENCE part 1) | Semaglutide | Adults with biopsy-proven metabolic dysfunction-associated steatohepatitis and fibrosis stage 2 or 3 | Semaglutide 2.4 mg once weekly | Placebo | Phase 3 histology trial | 1195 | Metabolic dysfunction-associated steatohepatitis |
| SELECT | Semaglutide | Adults aged 45 years or older with overweight or obesity and established cardiovascular disease, without diabetes | Semaglutide 2.4 mg once weekly | Placebo | Event-driven cardiovascular outcomes trial | 17604 | Established cardiovascular disease |
| SUSTAIN-6 | Semaglutide | Adults with type 2 diabetes mellitus at high cardiovascular risk | Semaglutide 0.5 mg and 1.0 mg once weekly | Placebo | 104 weeks | 3297 | Type 2 diabetes mellitus / cardiovascular risk |
| FLOW | Semaglutide | Adults with type 2 diabetes mellitus and chronic kidney disease | Semaglutide 1.0 mg once weekly | Placebo | Event-driven kidney outcomes trial | 3533 | Chronic kidney disease |
| SUSTAIN-1 | Semaglutide | Adults with type 2 diabetes mellitus inadequately controlled with diet and exercise alone | Semaglutide 0.5 mg and 1.0 mg once weekly | Placebo | 30 weeks | 387 | Type 2 diabetes mellitus |
| SUSTAIN-5 | Semaglutide | Adults with type 2 diabetes mellitus inadequately controlled on basal insulin | Semaglutide 0.5 mg and 1.0 mg once weekly | Placebo | 30 weeks | 396 | Type 2 diabetes mellitus |
| SUSTAIN-9 | Semaglutide | Adults with type 2 diabetes mellitus on sodium-glucose cotransporter-2 inhibitor background therapy | Semaglutide 1.0 mg once weekly | Placebo | 30 weeks | 302 | Type 2 diabetes mellitus |
| SURMOUNT-1 / prediabetes | Tirzepatide | Adults with obesity or overweight without diabetes; subgroup includes prediabetes | Tirzepatide 5 mg, 10 mg, 15 mg once weekly | Placebo | 72 weeks | 1032 | Obesity/overweight |
| SURMOUNT-2 | Tirzepatide | Adults with obesity or overweight and type 2 diabetes mellitus | Tirzepatide 10 mg and 15 mg once weekly | Placebo | 72 weeks | 938 | Obesity/overweight + type 2 diabetes mellitus |
| SURMOUNT-3 | Tirzepatide | Adults with obesity or overweight without diabetes after intensive lifestyle run-in | Tirzepatide maximum tolerated dose (10 mg or 15 mg) once weekly | Placebo | 72 weeks | 579 | Obesity/overweight |
| SURMOUNT-CN | Tirzepatide | Chinese adults with obesity or overweight and weight-related complications | Tirzepatide 10 mg and 15 mg once weekly | Placebo | 52 weeks | 210 | Obesity/overweight |
| SURMOUNT-J | Tirzepatide | Japanese adults with obesity disease | Tirzepatide 5 mg, 10 mg, 15 mg | Placebo | 72 weeks | 225 | Obesity/overweight |

| | | | | | | | |
|---|---|---|---|---|---|---|---|
| | | | once weekly | | | | |
| SURMOUNT-OSA trial 1 | Tirzepatide | Adults with moderate-to-severe obstructive sleep apnea and obesity, not using positive airway pressure | Tirzepatide 10 mg or 15 mg once weekly | Placebo | 52 weeks | 234 | Obesity + obstructive sleep apnea |
| SURMOUNT-OSA trial 2 | Tirzepatide | Adults with moderate-to-severe obstructive sleep apnea and obesity, using positive airway pressure | Tirzepatide 10 mg or 15 mg once weekly | Placebo | 52 weeks | 233 | Obesity + obstructive sleep apnea |
| SURPASS-1 | Tirzepatide | Adults with type 2 diabetes mellitus inadequately controlled with diet and exercise alone | Tirzepatide 5 mg, 10 mg, 15 mg once weekly | Placebo | 40 weeks | 478 | Type 2 diabetes mellitus |
| SURPASS-5 | Tirzepatide | Adults with type 2 diabetes mellitus inadequately controlled on insulin glargine | Tirzepatide 5 mg, 10 mg, 15 mg once weekly | Placebo | 40 weeks | 475 | Type 2 diabetes mellitus |
| SURPASS-CN-INS | Tirzepatide | Chinese adults with type 2 diabetes mellitus on basal insulin | Tirzepatide 10 mg once weekly | Placebo | Placebo-controlled phase 3 trial | 257 | Type 2 diabetes mellitus |

Table 1: Characteristics of included studies

### Acute Pancreatitis with GLP-1s vs Placebo

Sensitivity Analysis — Fixed-effect Mantel–Haenszel Risk Ratios with 95% CI

Semaglutide  Tirzepatide  Pooled estimate

| Study | Drug events / total | Placebo events / total | Risk Ratio (95% CI) | Weight (%) |
|---|---|---|---|---|
| STEP 1 | 3 / 1306 | 0 / 655 | 3.51 [0.18, 67.92] | 1.70 |
| STEP 2 | 1 / 807 | 1 / 403 | 0.50 [0.03, 7.96] | 1.94 |
| STEP 3 | 0 / 407 | 0 / 204 | Not estimable | — |
| STEP 5 | 0 / 152 | 0 / 152 | Not estimable | — |
| STEP 6 | 0 / 299 | 0 / 101 | Not estimable | — |
| STEP 7 | 0 / 249 | 0 / 126 | Not estimable | — |
| STEP 8 | 0 / 126 | 0 / 85 | Not estimable | — |
| STEP 9 | 0 / 271 | 0 / 136 | Not estimable | — |
| STEP 10 | 2 / 138 | 0 / 69 | 2.52 [0.12, 51.74] | 1.63 |
| STEP 11 | 0 / 101 | 0 / 49 | Not estimable | — |
| STEP-HFpEF | 0 / 263 | 1 / 266 | 0.34 [0.01, 8.24] | 1.46 |
| STEP-HFpEF DM | 2 / 310 | 0 / 306 | 4.94 [0.24, 102.39] | 1.62 |
| STEP UP | 3 / 1205 | 0 / 201 | 1.17 [0.06, 22.61] | 1.70 |
| STEP UP T2D | 1 / 410 | 0 / 102 | 0.75 [0.03, 18.32] | 1.46 |
| MASH phase 3 | 3 / 800 | 2 / 395 | 0.74 [0.12, 4.41] | 4.67 |
| SELECT | 17 / 8803 | 24 / 8801 | 0.71 [0.38, 1.32] | 38.65 |
| SUSTAIN-6 | 9 / 1648 | 12 / 1649 | 0.75 [0.32, 1.78] | 20.06 |
| FLOW | 10 / 1767 | 7 / 1766 | 1.43 [0.54, 3.74] | 16.03 |
| SUSTAIN-1 | 0 / 258 | 0 / 129 | Not estimable | — |
| SUSTAIN-5 | 0 / 263 | 0 / 133 | Not estimable | — |
| SUSTAIN-9 | 0 / 151 | 0 / 151 | Not estimable | — |
| SURMOUNT-1 / prediabetes | 3 / 762 | 1 / 270 | 1.06 [0.11, 10.09] | 3.44 |
| SURMOUNT-2 | 2 / 623 | 1 / 315 | 1.01 [0.09, 11.10] | 3.67 |
| SURMOUNT-3 | 1 / 287 | 1 / 292 | 1.02 [0.06, 16.35] | 1.97 |
| SURMOUNT-CN | 0 / 141 | 0 / 69 | Not estimable | — |
| SURMOUNT-J | 0 / 150 | 0 / 75 | Not estimable | — |
| SURMOUNT-OSA trial 1 | 0 / 114 | 0 / 120 | Not estimable | — |
| SURMOUNT-OSA trial 2 | 2 / 119 | 0 / 114 | 4.90 [0.23, 103.14] | 1.66 |
| SURPASS-1 | 0 / 363 | 0 / 115 | Not estimable | — |
| SURPASS-5 | 0 / 355 | 0 / 120 | Not estimable | — |
| SURPASS-CN-INS | 0 / 193 | 0 / 64 | Not estimable | — |
| **Total (fixed-effect MH)** | **59 / 22841** | **50 / 17433** | **0.99 [0.67, 1.46]** | **100.0** |

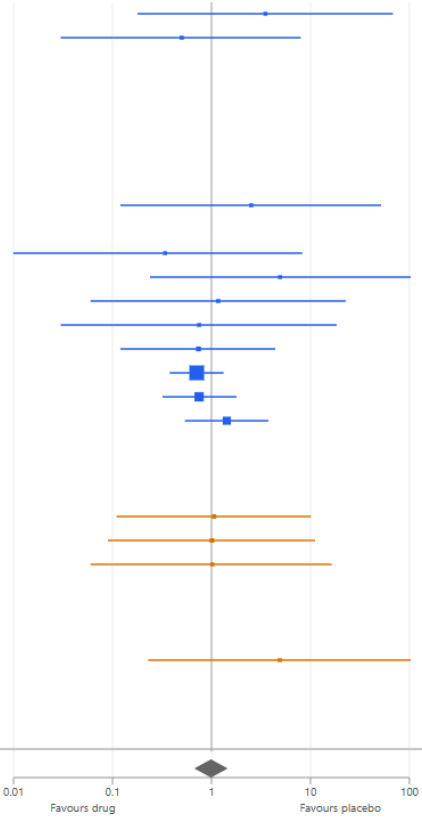


**Model:** Fixed-effect Mantel–Haenszel risk ratio | **Overall effect:** Z = 0.07, p = 0.95 | **Heterogeneity:** $Chi^2$ = 5.92, df = 14, p = 0.97 | **Inconsistency:** $I^2$ = 0%

Supplementary Figure 1: Sensitivity analysis for risk ratio

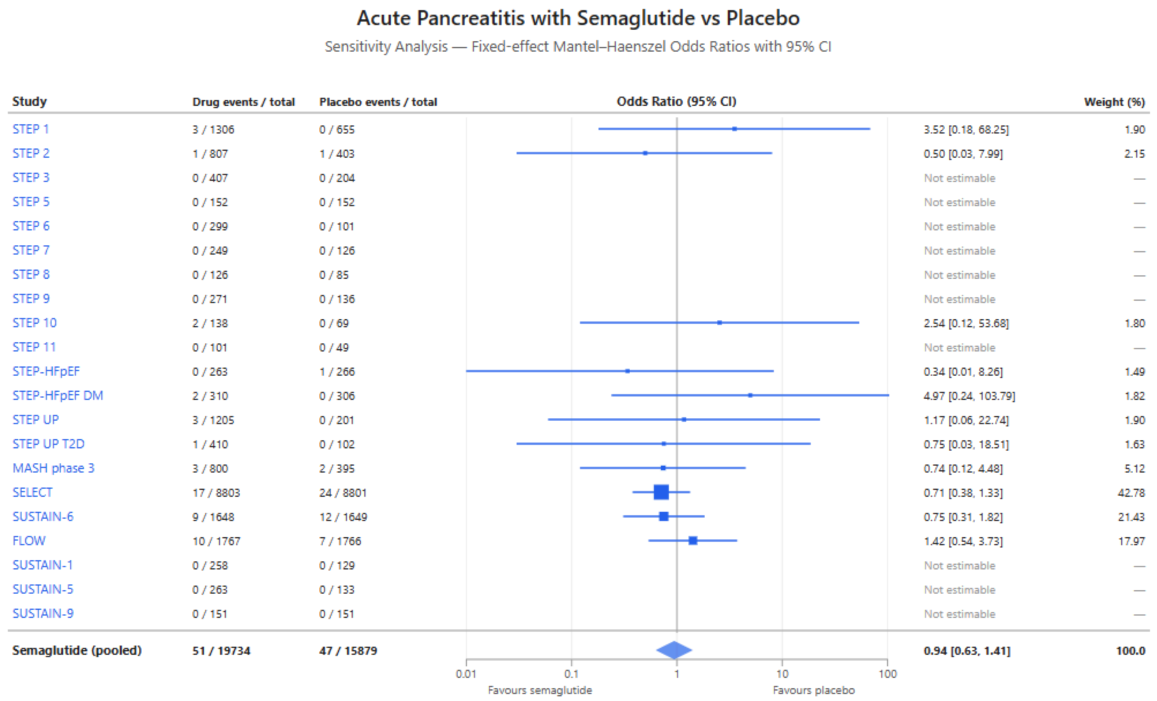


**Model:** Fixed-effect Mantel–Haenszel | **Overall effect:** Z = 0.29, p = 0.77 | **Heterogeneity:** Chi² = 4.71, df = 10, p = 0.91 | **Inconsistency:** I² = 0%

Supplementary Figure 2: Subgroup analysis for semaglutide

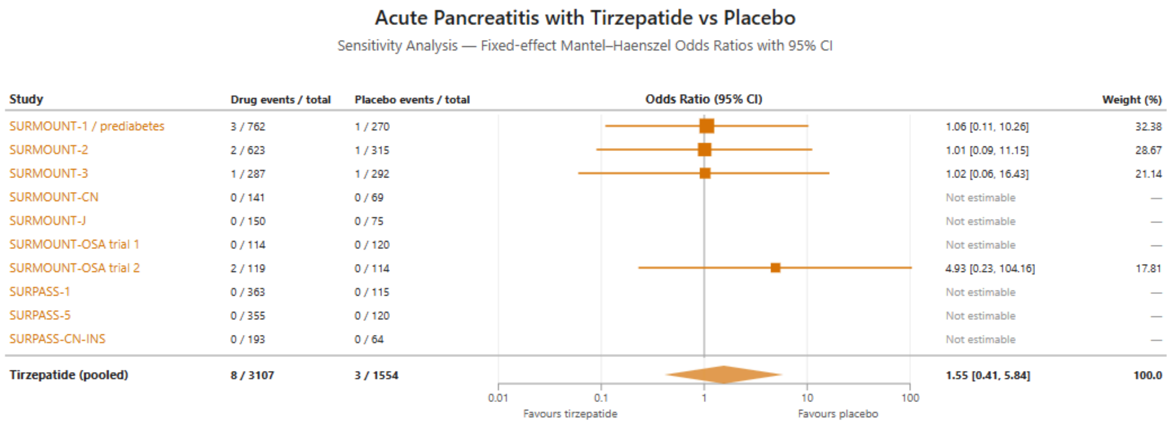


**Model:** Fixed-effect Mantel–Haenszel | **Overall effect:** Z = 0.65, p = 0.52 | **Heterogeneity:** Chi² = 0.86, df = 3, p = 0.83 | **Inconsistency:** I² = 0%

Supplementary Figure 3: Subgroup analysis for tirzepatide

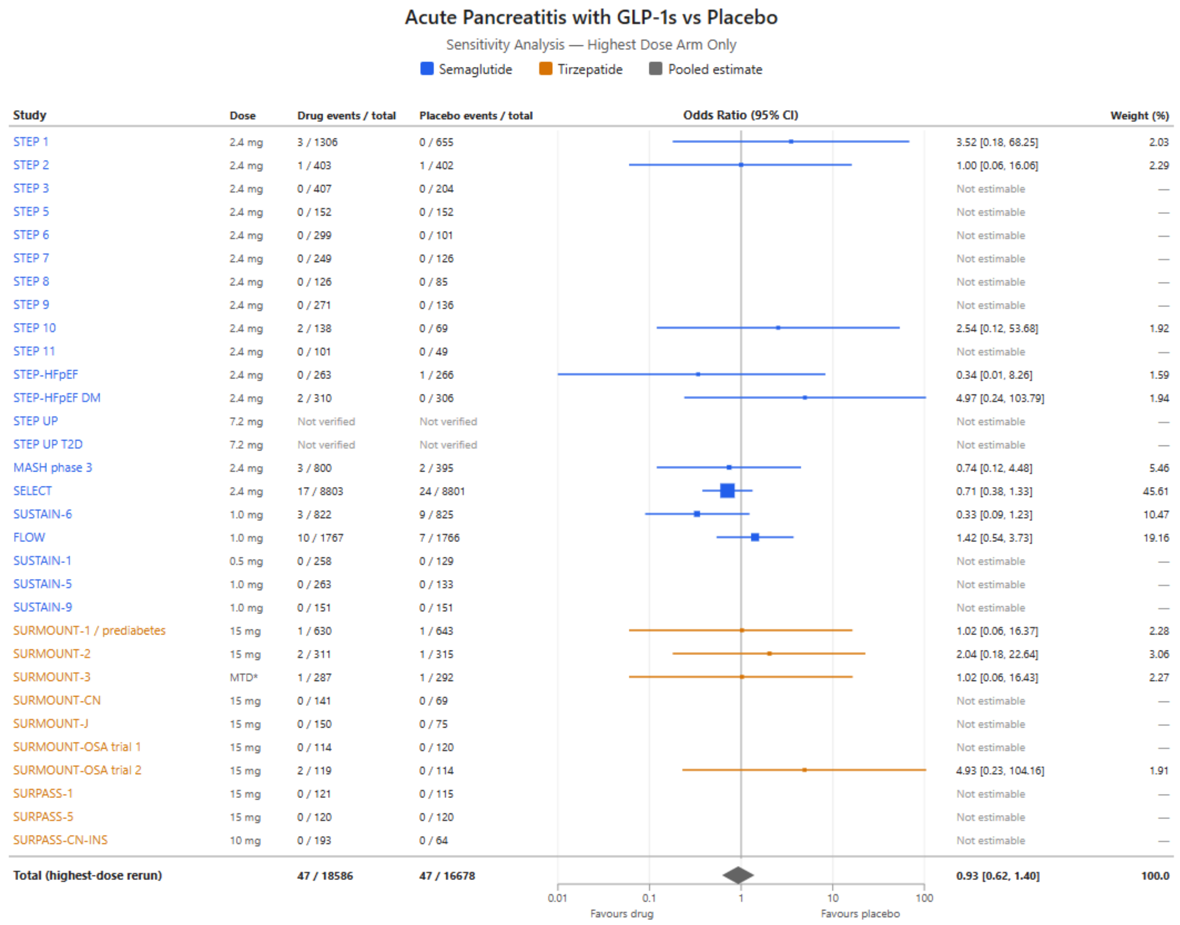


Supplementary Figure 4: Subgroup analysis by dose

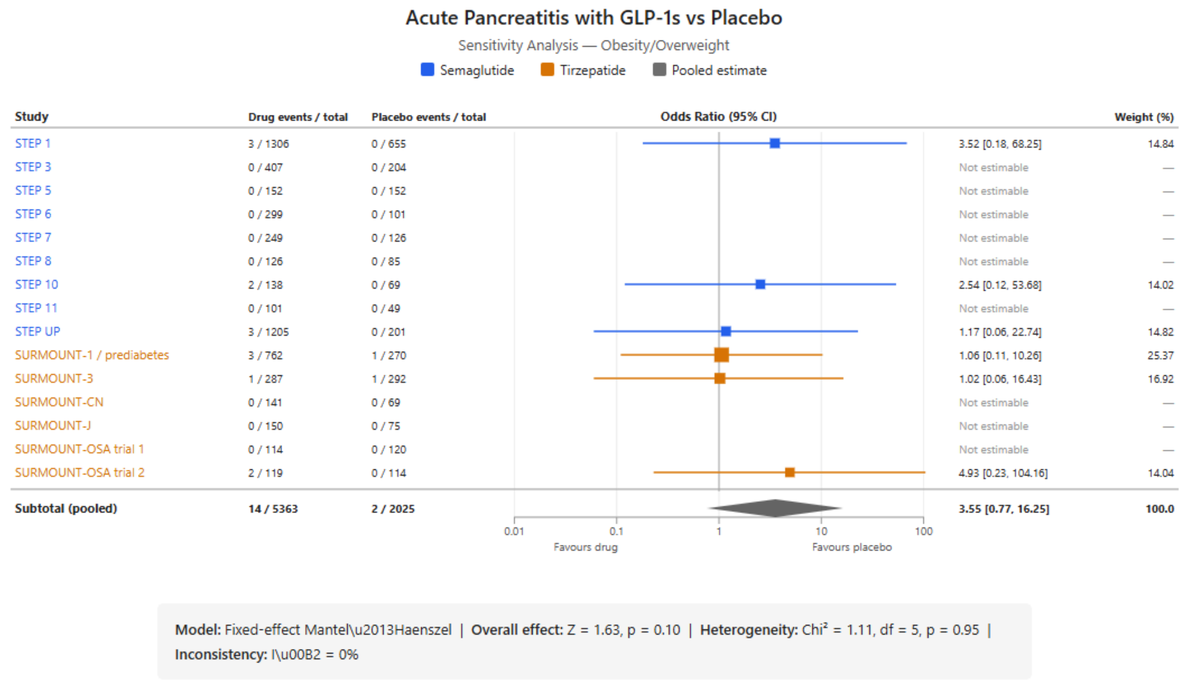


Supplementary Figure 5: Subgroup analysis by overweight/obesity

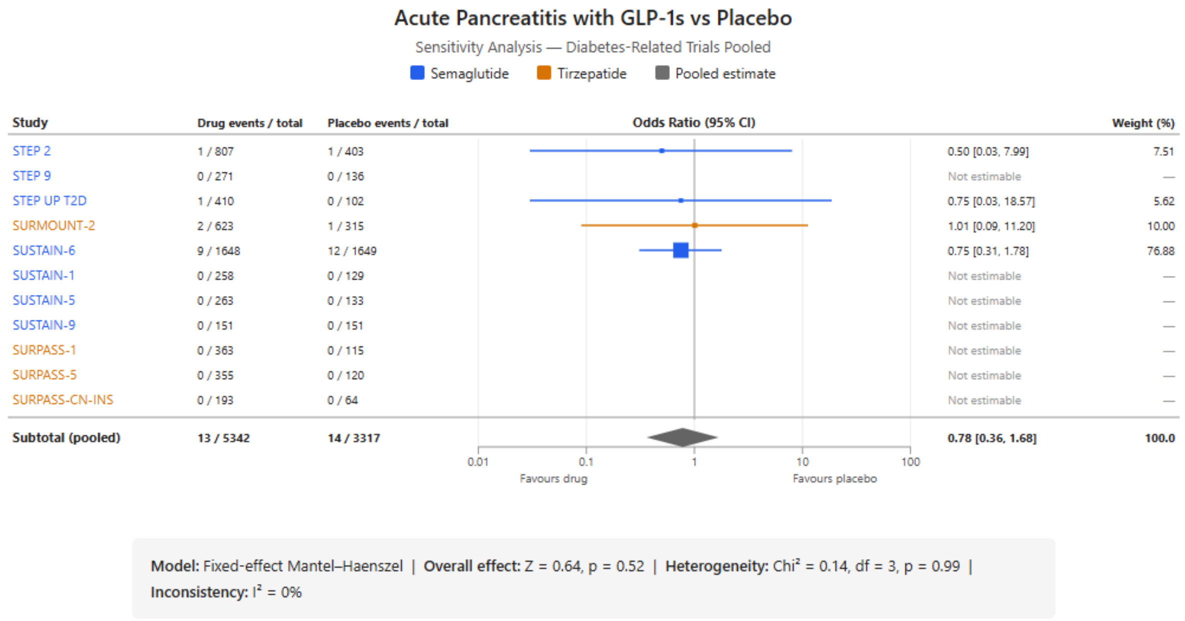


Supplementary Figure 6: Subgroup analysis by type 2 diabetes

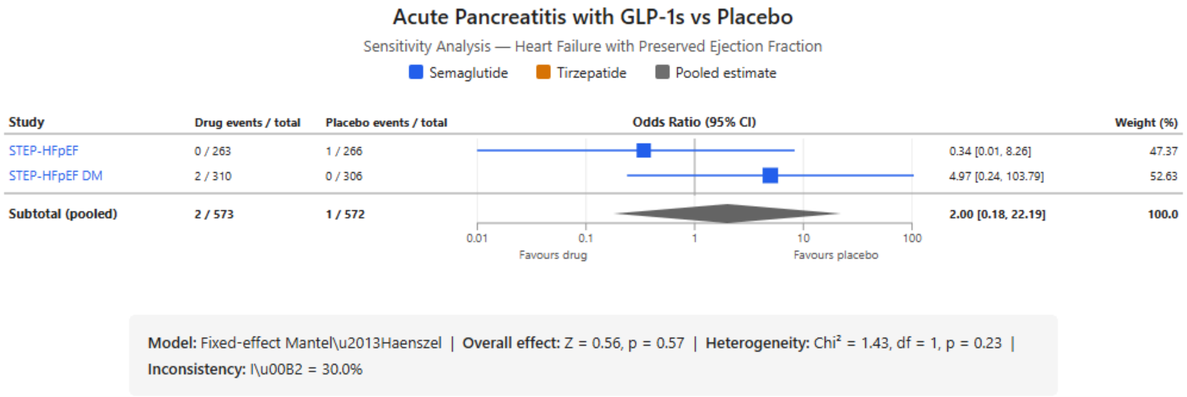


Supplementary Figure 7: Subgroup analysis by heart failure

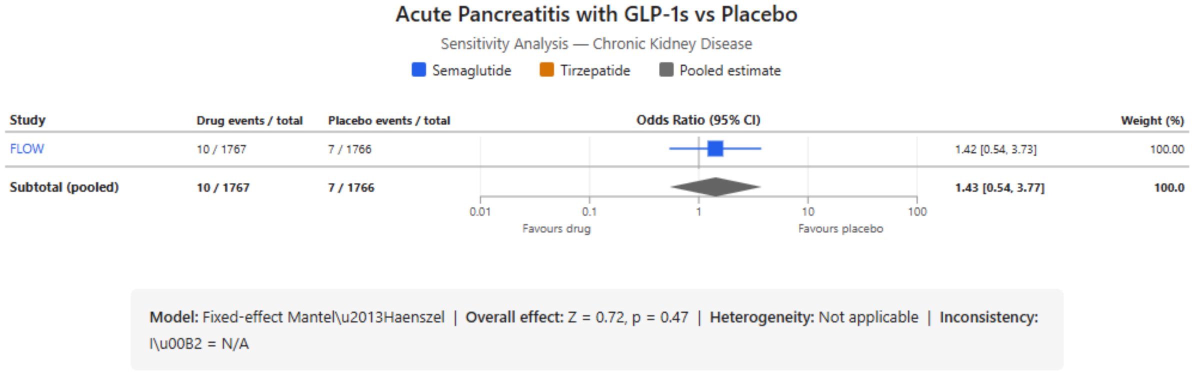


Supplementary Figure 7: Subgroup analysis by chronic kidney disease

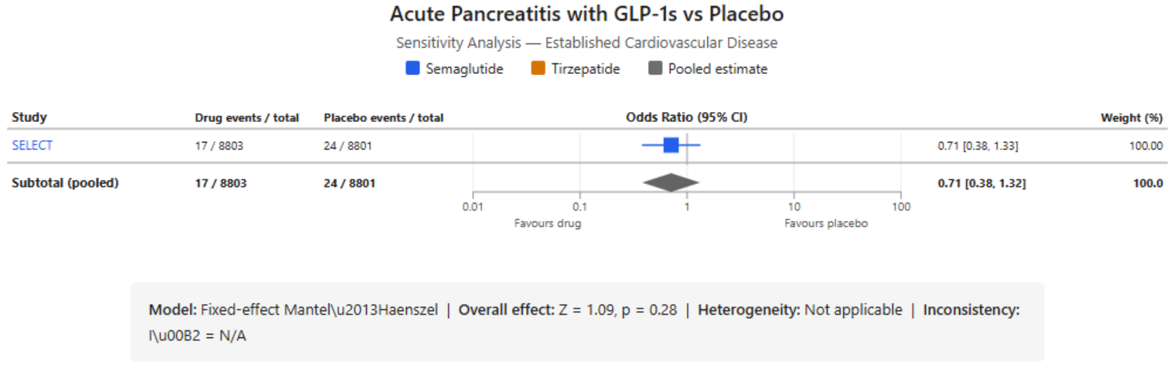


Supplementary Figure 8: Subgroup analysis by cardiovascular disease

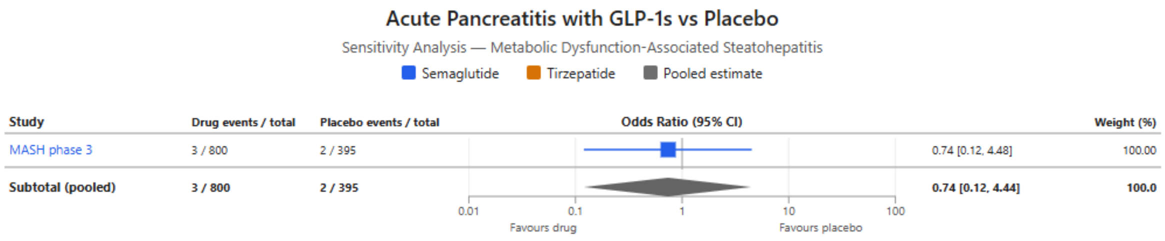


**Model:** Fixed-effect Mantel\u2013Haenszel | **Overall effect:** Z = 0.33, p = 0.74 | **Heterogeneity:** Not applicable | **Inconsistency:** I\u00B2 = N/A

Supplementary Figure 9: Subgroup analysis by metabolic dysfunction-associated steatohepatitis

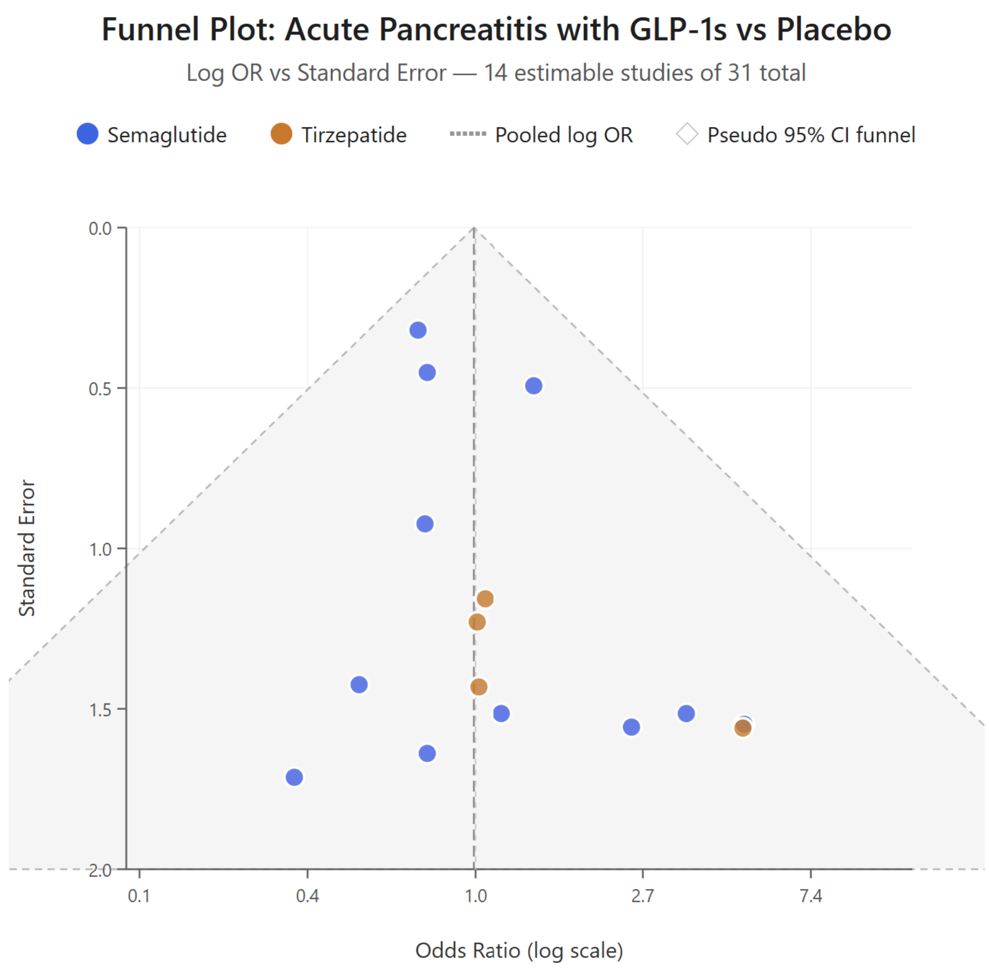


#### Formal Tests of Funnel Plot Asymmetry

| | |
|---|---|
| **Egger's regression test** | intercept = 0.49 (SE 0.27), $t$ = 1.80, df = 13, $p$ = 0.095 |
| **Begg's rank correlation test** | Kendall's $\tau$ = 0.23, $z$ = 1.14, $p$ = 0.255 |

Supplementary Figure 9: Publication bias funnel plot
